# A quasi-experimental evaluation of a financial incentive for first-dose COVID-19 vaccination among adults aged ≥ 60 years in South Africa

**DOI:** 10.1101/2022.05.06.22274712

**Authors:** Candice M Chetty-Makkan, Harsha Thirumurthy, Elizabeth F Bair, Simamkele Bokolo, Candy Day, Korstiaan Wapenaar, Jesse Werner, Lawrence Long, Brendan Maughan-Brown, Jacqui Miot, Sophie J S Pascoe, Alison M Buttenheim

**Affiliations:** Health Economics and Epidemiology Research Office, Faculty of Health Sciences, University of Witwatersrand, Johannesburg, South Africa; Department of Medical Ethics and Health Policy, Perelman School of Medicine, University of Pennsylvania, Philadelphia, PA, USA; DG Murray Trust, Cape Town, South Africa; Genesis Analytics, Johannesburg, South Africa; Department of Global Health, Boston University School of Public Health, Boston, MA, USA; University of Cape Town, Southern Africa Labour and Development Research Unit, South Africa; Department of Family and Community Health, School of Nursing, University of Pennsylvania, Philadelphia PA, USA

## Abstract

**Introduction:** COVID-19 vaccination coverage in South Africa (RSA) remains low despite increased access to vaccines. On November 1, 2021, RSA introduced the Vooma Voucher program which provided a small guaranteed financial incentive, a Vooma Voucher redeemable at grocery stores, for COVID-19 vaccination among older adults, a population most vulnerable to serious illness, hospitalization, and death. However, the association of financial incentives with vaccination coverage remains unclear.

**Methods:** We evaluated the association of the conditional economic incentive program with first-dose vaccination rates among adults (aged ≥60 years) through a quasi-experimental cohort study. The Vooma Voucher program was a nationwide vaccination incentive program implemented for adults aged ≥60 years from November 1, 2021 to February 28, 2022. We ran interrupted time series models to evaluate the Vooma Voucher program at national and provincial levels. We used data between October 1, 2021 and November 27, 2021 in models estimated at the daily level. Individuals who received their first vaccine dose received a text message to access a ZAR100 ($∼7) voucher that was redeemable at grocery stores.

**Results:** The Vooma Voucher program was associated with a 7.15-12.01% increase in daily first-dose vaccinations in November 2021 compared to late October 2021. Overall, the incentive accounted for 6,476-10,874 additional first vaccine doses from November 1-27, 2021, or 8.31-13.95% of all doses administered to those aged ≥60 years during that period. This result is robust to the inclusion of controls for the number of active vaccine delivery sites and for the nationwide Vooma vaccination weekend initiative (November 12-14), both of which also increased vaccinations through expanded access to vaccines and demand creation activities.

**Conclusions:** Financial incentives for COVID-19 vaccination led to a modest increase in first dose vaccinations among older adults in RSA. Financial incentives and expanded access to vaccines may result in higher vaccination coverage.

**Trial registration number (SANCTR):** DOH-27-012022-9116

**Key points (3-5 sentences):** *What is already known about this topic?:* There is a lack of evidence on whether financial incentives for COVID-19 vaccinations are effective in low- and middle-income countries.

*What does this study add?:* We found that a ZAR100 (∼US$7) incentive for adults aged ≥60 years increased additional first vaccine doses between November 1-27, 2021 to those aged ≥60 years during that period.

*How this study might affect research, practice or policy?:* Small guaranteed financial incentives may be an effective strategy to increase vaccine demand among older adults in low- and middle-income countries.

## Introduction

COVID-19 infections in sub-Saharan Africa (SSA) were high when compared to many other countries due to the highly transmissible Delta variant (1,2). South Africa recorded its first COVID-19 case on March 5th, 2020 and enforced a strict lockdown and other mitigation measures (3). These restrictions, while effective, were insufficient to prevent multiple waves of COVID-19 infections (4) with significant morbidity and mortality (1).

South Africa had one of the earliest and most robust vaccination programs on the continent. The South African National Department of Health (NDoH) ensured equitable access to free COVID-19 vaccination services (5,6) through a network of vaccination sites and stimulated demand through mass media campaigns. In South Africa, COVID-19 vaccines were widely available for individuals aged ≥60 years by May 2021 (1,2). Despite robust efforts to promote vaccines and ensure easy access, by October 2021 only 63% of older adults had received at least one vaccine dose, well below the NDoH targets to protect vulnerable populations (2).

As in other countries, there was substantial interest for financial incentives to increase vaccine uptake. Financial incentives increase the immediate benefits of vaccination, and can mitigate perceived costs of and barriers to vaccination, including hesitancy related to vaccine safety and hassle factors associated with accessing vaccination services (7). Evidence on the effectiveness of incentives for COVID-19 vaccination comes almost exclusively from high-income countries and is mixed (8–15): some studies found that guaranteed rewards can increase vaccination (8,9), while others showed no effect of guaranteed or lottery incentives (10,16,17), or indicated that incentives may backfire by decreasing vaccination intentions (11). Low- and middle -income countries (LMICs), have faced a substantial burden of COVID-19, but there is lack of evidence on whether incentives for COVID-19 are effective in LMICs.

In South Africa, like other African regions, low levels of education among the elderly and limited competence with digital technologies were barriers to accessing COVID-19 vaccination services (3). Around 73% of people over the age of 60 live off South Africa’s old age pension grant that is approximately ZAR 1980 (∼$139) per month (3,4). In order to increase inclusion of the elderly into the national-level COVID-19 response plan, the Vooma Voucher program was one opportunity to mitigate the financial constraints experienced by the elderly to access vaccination services, findings of which can be generalised to other LMICs.

To increase vaccination among older adults in advance of an anticipated COVID-19 fourth wave, the NDoH Vooma Voucher program was launched on November 1, 2021 to reach the vulnerable and impoverished. The voucher program was promoted through multiple communication channels (eFigures) and continued until February 28, 2022, with some changes in eligibility and voucher amounts (eMethods). These approaches included use of the Electronic Vaccination Data System (EVDS) to send invites to any individual who was eligible to receive the vaccine. There were also social media posts on National Department of Health (NDoH) channels (WhatsApp groups, Facebook, Instagram, and Twitter, the RSA coronavirus website, FAQ Knowledge base (which is a helpdesk system linked to the RSA coronavirus website), other media (press briefings, media statements by the Minister of Health) and radio public service announcements. In addition, awareness of the Vooma Voucher program took place through the network of district communicators, SMSs via the Government Communication and Information System (GCIS) and secondary promotion through general media stories (2). These communication platforms were extensive and vast when compared to other settings where targeted recruitment methods were used (12,17).

The Vooma Voucher program was initially available to adults aged ≥60 years. Individuals who received their first vaccine dose received a ZAR100 (∼$7) voucher by text message that was redeemable at local grocery stores. For many individuals ≥60 years, their monthly income will come from the Old Age Pension (4). For those individuals receiving the Old Age Pension (i.e. individuals at the bottom of the income distribution in this age range) the voucher value was equivalent to approximately 1.5 days of income. The purpose of the voucher was to compensate a person for the travel and costs that might be incurred by going for vaccination. The voucher value was intended to reduce financial barriers to vaccination while not being so high as to increase the risk of undue influence. Relative to local income levels, the incentive amount in South Africa was higher than the amounts offered in several studies in the US and Europe (17).

We assessed the short-term association of the Vooma Voucher incentive program on first-dose vaccination rates among those aged ≥60 years.

## Methods

The South African population is 60.1 million comprising 81% black South Africans, with Gauteng and KwaZulu-Natal provinces accounting for 45.4% of the population. In the Gauteng and KwaZulu-Natal provinces, the proportion of those ≥60 years is 8.5% and 8.2% respectively. Gauteng also has the highest net inflow of migrants (18). During the COVID-19 pandemic waves, the crude death rate (CDR) within a year increased from 8,7 deaths per 1000 in 2020 to 11,6 deaths per 1000 people in 2021. According to the Medical Research Council (MRC) weekly reports, there were more than 180 000 excess deaths in South Africa since March 2020 (18).

We used national data on the number of first doses of the COVID-19 vaccine that were administered daily. The analysis was based on data provided by the NDoH for purposes of evaluating the incentive program. Using population data from the South African Community Survey 2016 (19) we calculated the number of first COVID-19 vaccine doses administered daily per 10,000 individuals aged ≥60 years. The Vooma Voucher programme was extended to those ≥ 50 years on November 18, 2021 and the voucher amount was increased to ZAR200 (US$14) on December 1, 2021. We did not include those 50-59 years in the analysis, as we only had data for 10 days on these individuals. A sensitivity analysis with weekly doses is in the eMethods. First dose vaccination rates were calculated separately at the national- and province-level.

### Patient and Public Involvement

For this analysis, deidentified, aggregate routine data from the National Department of Health COVID-19 Vaccination program was used. Results from this analysis will be disseminated through policy briefs, social media platforms and published reports.

## Data analysis

In the absence of a randomised control trial, we selected the most rigorous study design in the form of a quasi-experimental method for this evaluation. Assumptions for the difference-in-differences approach were violated, therefore we did not use this model. Instead, we estimated four separate interrupted time series (ITS) models to estimate the association between the Vooma Voucher programs and trends in vaccination rates: unadjusted and adjusted national models as well as unadjusted and adjusted provincial models. The primary outcome was the daily vaccination rate per 10,000 individuals aged ≥60 years in South Africa. National models utilized linear regression with Newey-West standard errors, while provincial models utilized generalized estimating equations with robust standard errors to account for province-level clustering of observations across time. Daily models included day of the week indicators to control for temporal patterns within each week and week-specific fixed effects. Unadjusted models included no additional covariates; adjusted models included additional measures of vaccine supply (daily active vaccine delivery sites per 100,000 individuals and an indicator variable for the three days of the Vooma Vaccination weekend). The Vooma Vaccination Weekend was a nationwide initiative to open more vaccination sites to boost demand (2).

Province-level adjusted models included an indicator for KwaZulu-Natal and Gauteng provinces, as well as an interaction term to assess the impact of the voucher program in these provinces versus the rest of the country. Gauteng (15.8M) and KwaZulu-Natal (11.5M) are South Africa’s two most populous provinces accounting for almost half (45.4%) of the country’s population (18). The vaccination coverage in these provinces was, and remains low, and addressing this gap was critical to the COVID response. The first dose vaccination coverage in the Gauteng (5.79 per 10,000) and KwaZulu-Natal (4.81 per 10,000) provinces were also much lower than the rest of the country (7.41 per 10,000) in the week before the introduction of the Vooma Voucher. Due to the population size and low vaccination rates in these areas, we selected these provinces for comparison to the rest of the country.

Our analysis included data from October 1, 2021 to November 27, 2021. Although the Vooma Voucher program continued until February 28, 2022, the announcement of the Omicron variant on November 24, 2022 would likely confound the effect of the incentive program on days that followed the announcement. Analyses were conducted using Stata 17.0 (StataCorp, College Station, TX). This study was approved by the University of Witwatersrand Human Research Ethics Committee (Medical) (211123).

## Results

In the weeks preceding introduction of the Vooma Voucher program, the 7-day rolling average of daily first vaccine doses administered declined from 13-15 doses per 10,000 individuals aged ≥60 years in early-September to <10 doses by late-October 2021 (**Figure 1**). For the first 27 days, the Vooma Voucher program was associated with an increase in daily first vaccine doses administered of 0.89 per 10,000 individuals (95% CI 0.52, 1.27; p<0.001) (**Table 1**); that equated to 10,874 additional doses compared to the number of first doses expected to be administered in the absence of the Vooma Voucher program. Adjusting for vaccine delivery sites and for the Vooma Vaccination Weekend reduced the increase associated with the Vooma Voucher program (+0.65 daily first doses per 10,000 individuals; 95% CI 0.33, 0.96; p<0.001, 7,942 additional doses). Taken together, these results suggest that 8.31-13.95% of the 77,947 first doses administered to individuals aged ≥60 years between November 1-27, 2021, may be attributed to the Vooma Voucher program (**Figure 2**). Using the last week of October 2021 as a reference, the percentage increase in vaccinations is estimated at 7.15% to 12.01% per 10, 000 individuals per day.

**Table 1:** Results from interrupted time-series analyses estimating the association of the Vooma voucher program with changes in the number of first vaccine doses delivered per day per 10,000 adults ≥60 years in South Africa.

|  | Unadjusted daily models |  | Adjusted daily models |  |
| --- | --- | --- | --- | --- |
|  | Coeff<br>(95% CI) | p-value | Coeff<br>(95% CI) | p-value |
| **National Model** |  |  |  |  |
| Vooma voucher program | 0.89<br>(0.52, 1.27) | p<0.001 | 0.65<br>(0.33, 0.96) | p<0.001 |
| Vooma vaccination weekend | -- | -- | 3.28<br>(1.98, 4.58) | p<0.001 |
| Daily active facilities per 100, 000 population | -- | -- | 0.55<br>(-0.60, 1.69) | p=0.340 |
| **Provincial Model** |  |  |  |  |
| Vooma voucher program | 0.80<br>(0.43, 1.18) | p<0.001 | 0.53<br>(0.17, 0.89) | p=0.004 |
| Vooma vaccination weekend | -- | -- | 2.10<br>(1.04, 3.16) | p<0.001 |
| Daily active facilities per 100, 000 population | -- | -- | 0.86<br>(0.43, 1.29) | p<0.001 |
| KwaZulu-Natal and Gauteng | -4.15<br>(-7.45, -1.58) | p=0.003 | -3.49<br>(-6.94, 0.04) | p=0.047 |
| Vooma voucher in KwaZulu- Natal and Gauteng | 1.96<br>(1.01, 2.91) | p<0.001 | 1.39<br>(0.34, 2.45) | p=0.010 |

Notes: Models also included day of the week and week fixed effects. In province-level models, we include an indicator for KwaZulu-Natal and Gauteng combined, compared to the rest of the country.

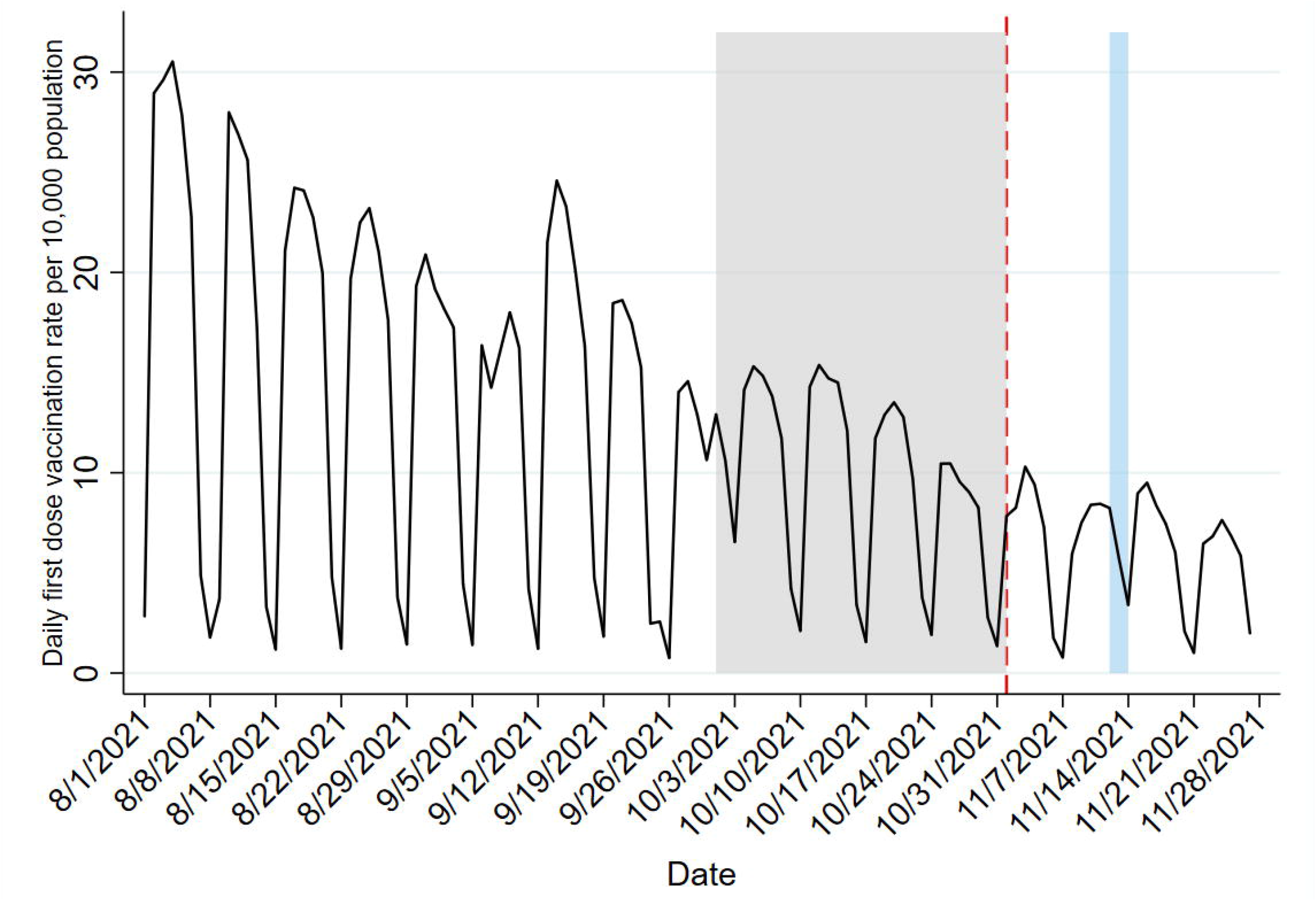

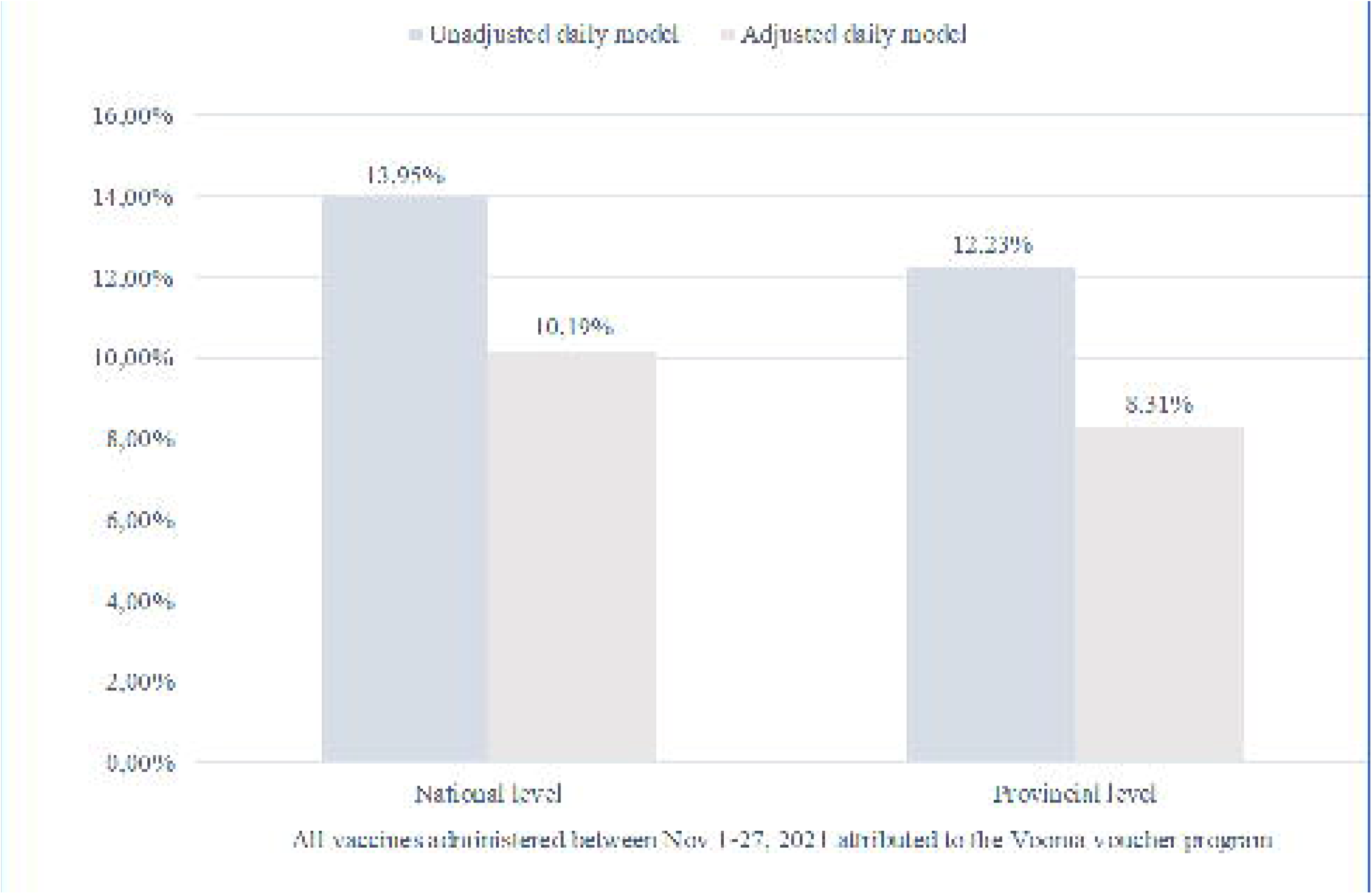

Provincial models showed directionally similar results (unadjusted: +0.80 daily first doses per 10,000 individuals; 95% CI 0.43, 1.18; p<0.001, 9,790 additional doses; adjusted: +0.53 daily first doses per 10,000 individuals; 95% CI 0.17, 0.89; p=0.004, 6,476 additional doses), **Table 1 and Figure 3**. The Vooma Voucher program was more effective in Gauteng and KwaZulu-Natal relative to other provinces (unadjusted: +1.96 daily first doses per 10,000 individuals; 95% CI 1.01-2.91; p<0.001; adjusted +1.39 daily first doses per 10,000 individuals; 95% CI 0.34-2.45; p=0.010). Weekly models and implications of supply and demand creation adjustments are found in the eTable.

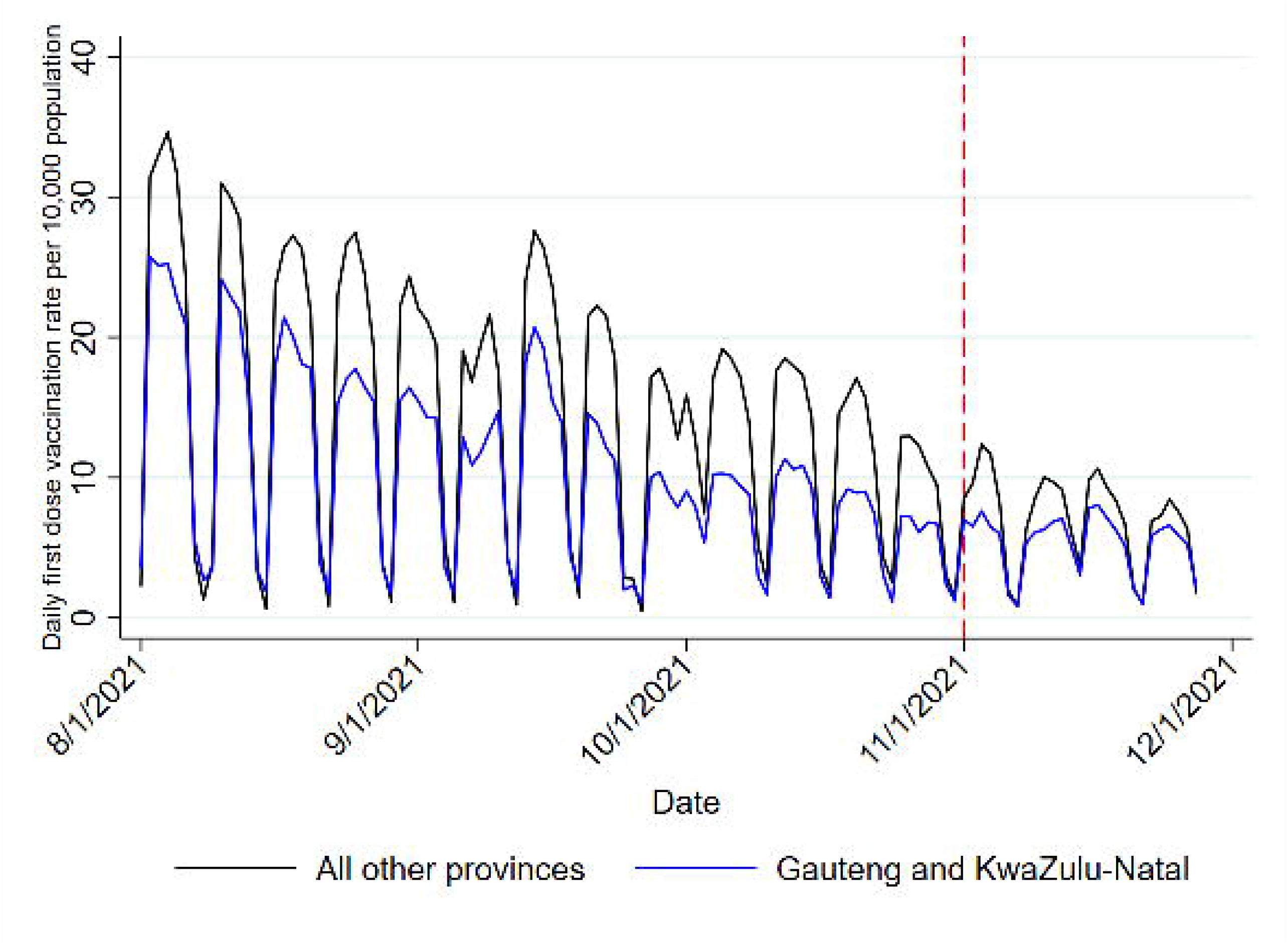

## Discussion

A nationwide financial incentive program that provided vouchers redeemable at grocery stores to older adults who received a first dose of a COVID-19 vaccine was associated with increased vaccinations in South Africa. In the first month of the program, before vaccine demand may have been influenced by the announcement of the Omicron variant, 8.31% to 13.95% of all first vaccine doses administered to older adults could be attributed to the program. This represents a meaningful increase in vaccinations in response to a ZAR100 (∼USD7) incentive.

This study is among the first to evaluate financial incentives for COVID-19 vaccination in LMICs. Evaluations of such incentives have primarily been conducted in high-income countries (8–10,16,17). An incentive of US$24 increased vaccination rates by 4% in a randomized trial in Sweden (8). Moreover, several studies found that lotteries and guaranteed incentives offered by US states did not increase vaccination rates (17). However, the findings from this evaluation show that providing small financial incentives to the elderly in resource limited settings can be one tool that boosts vaccine uptake. These findings contribute to the limited body of literature on financial incentives and COVID-19 vaccine uptake and provide the groundwork for further analyses that could inform policy changes.

Our results also show that the Vooma Voucher program was able to increase first dose vaccine uptake in specific provinces such as Gauteng and KwaZulu-Natal relative to other provinces. Due to the high population and high mobility of individuals within Gauteng and KwaZulu-Natal, it is possible that there was a higher uptake of the Vooma Vouchers when compared to other provinces. Although there was some mistrust in the community during the roll-out of the COVID-19 vaccine program, it is plausible that endorsement of the Vooma Voucher program (20) by key figures and Vooma Vaccination Weekends through various platforms were associated with higher vaccination rates.

The Vooma Voucher may have had better results due to substantial advertisement of the program and the amount of the incentive relative to income level of the target population in South Africa. From our evaluation it is helpful to know that the Vooma Voucher of ZAR100 ($7) is one potentially effective tool in the toolkit, however we will need all costs for the program in order to be able to draw any conclusions regarding the feasibility and scalability of this program. Given the available data we are only able to speak directly to its effectiveness.

A key limitation is the assumption in our ITS models that there were no other factors that coincided with the introduction of the Vooma Voucher program and affected vaccine demand. The robustness of our findings to the inclusion of supply and demand creation measures, and to provincial and weekly specifications, increases confidence in our findings. Another limitation is that we only study short-term effects of the incentive program, as the announcement of the Omicron variant made it challenging to study effects of incentives beyond that date using ITS analysis. We also did not control for infection rate due to the low testing rate in this context. According to the updated prioritized COVID-19 testing guidance, hospitalised patients, persons with symptoms of COVID-19 infection and individuals who were in close contact with confirmed cases including asymptomatic contacts were eligible for testing (21). This limited testing showed that the general population did not have access to widespread testing. There were also constraints on testing during the waves. Therefore, it is not clear that infection rate was a major driver of population level behaviour.

Given the vulnerability of older adults to serious illness, hospitalization, and death as a result of COVID-19, identifying effective strategies to increase vaccine demand is crucial. More generally, as LMICs struggle to achieve sufficiently high vaccine demand despite expansions in vaccine delivery and access, our findings suggest that small financial incentives may be effective in increasing vaccination coverage.

## Data Availability

All data produced in the present study are available upon reasonable request to the authors

## Author Contributions

**Chetty-Makkan, Thirumurthy, Bair, Bokolo, Day, Wapenaar, Werner & Buttenheim** had full access to all the data in the study and take responsibility for the integrity of the data and the accuracy of the data analysis.

**Concept and design:** Chetty-Makkan, Thirumurthy, Bair, Bokolo & Buttenheim

**Drafting of the manuscript:** Chetty-Makkan, Thirumurthy, Bair, Bokolo & Buttenheim

**Critical revision of the manuscript for important intellectual content:** Thirumurthy, Long, Maughan-Brown, Miot, Pascoe, Buttenheim

**Statistical analysis:** Thirumurthy, Bair and Buttenheim

**Obtained funding:** Chetty-Makkan, Maughan-Brown, Pascoe, Miot & Buttenheim

**Administrative, technical, or material support:** Chetty-Makkan, Bair, Bokolo, Day, Wapenaar, Werner, Buttenheim

**Supervision:** Thirumurthy & Buttenheim

## Acknowledgments

We would like to thank Dr Zameer Brey, from the Bill and Melinda Gates Foundation, and Mr Gaurang Tanna, formerly from the National Department of Health, for their technical support and guidance in this study. We would also like to thank the National Department of Health COVID-19 Vaccination Management Team for access to data.

