## Supplementary material for "A quasi-experimental evaluation of a financial incentive for first-dose COVID-19 vaccination among adults aged ≥ 60 years in South Africa": Online supplement

### Online- only Material

**eFigure:** Vooma Voucher Promotion Pack for individuals ≥ 60 years old

**eFigure:** Average daily first dose vaccination rate per 10,000 adults ≥60 years in South Africa by week, for the weeks of August 16, 2021 through November 22, 2021

**eMethods:** Further details on Vooma Voucher program analysis

**eTable:** Results from interrupted time-series analyses estimating the association of the Vooma voucher program with changes in the number of first vaccine doses delivered per week per 10,000 adults ≥60 years in South Africa

**eFigure: Vooma Voucher Promotion Pack for individuals ≥ 60 years old**

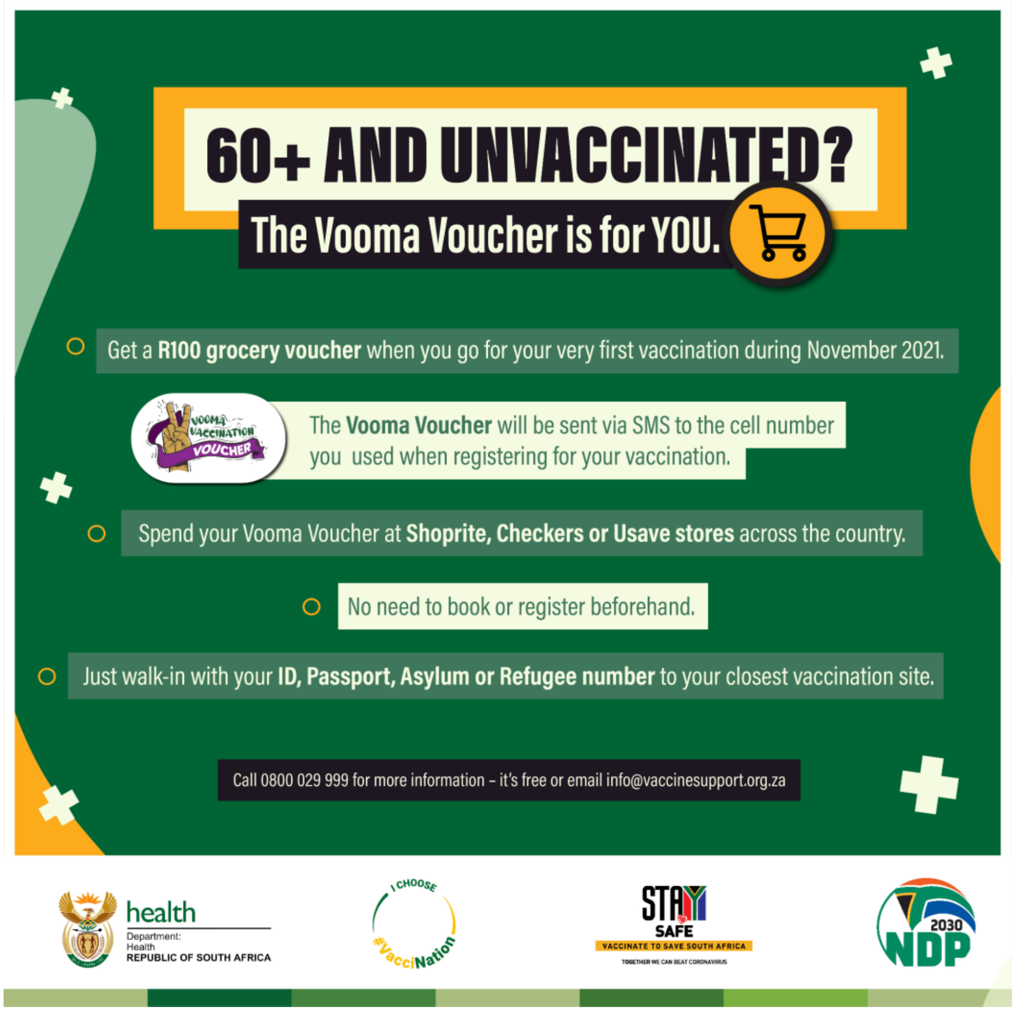


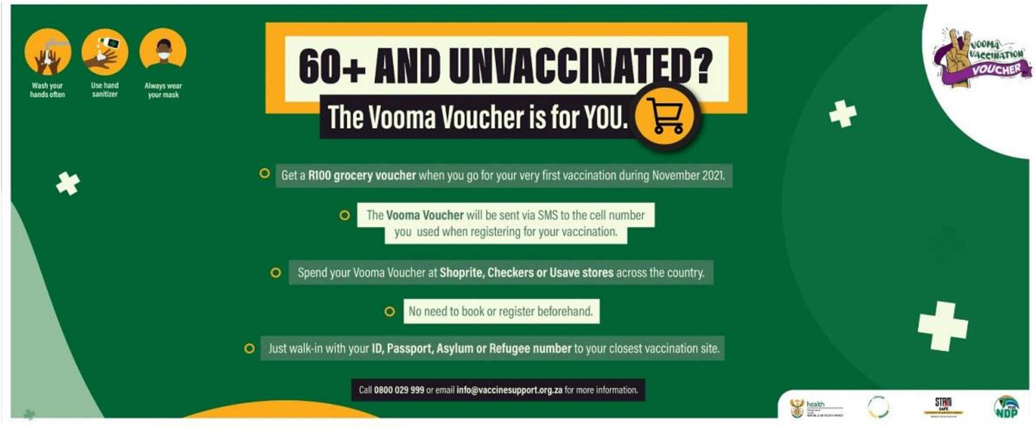


Reference: Resources are from the DG Murray Trust

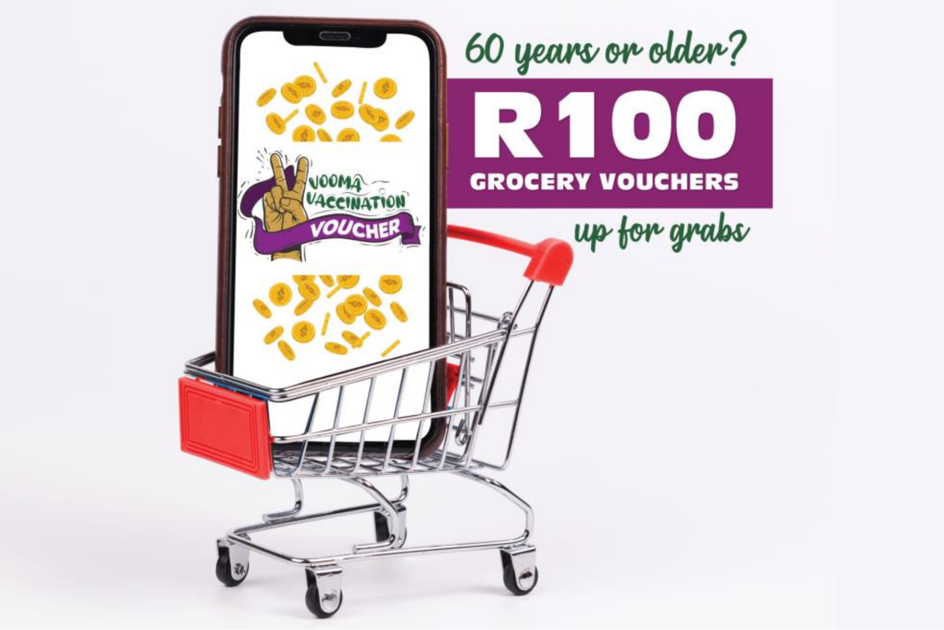


Reference: Mathieson, 2021

**eMethods:**

Vooma Voucher Program:

Eligibility was confirmed and the voucher notification was sent automatically through South Africa's national Electronic Vaccination Data System (EVDS), a self-registration application used in the COVID-19 national vaccine programme. Vouchers were valid for 60 days after receipt and could be redeemed at a local grocery store. The programme was extended to those ≥ 50 years on November 18, 2021 and the voucher amount was increased to ZAR200 (US$14) on December 1, 2021.

Additional details on daily models:

Based on visual inspection, we began pre-intervention data for the daily models on October 1st, 2021. In adjusted daily models we included vaccination facilities active per 100,000 population (at the national- or provincial-level, as appropriate) to control for vaccine supply, as well as an indicator variable for the Vooma Weekend (November 12-14, 2021), a nationwide initiative of focused demand creation, vaccination supply, and transport support initiatives. Visual inspection as a technique is an accepted approach used in ITS to help determine the appropriate pre-intervention window (Bernal, Cummins, and Gasparrini, 2017. In province-level adjusted models we also included an indicator for the provinces of KwaZulu-Natal and Gauteng compared to the rest of the country, as well as an interaction term for the impact of the program in KwaZulu-Natal and Gauteng compared to the rest of the country. This second term is meant to look for heterogeneous treatment effects in the two sets of provinces.

In all models (daily and weekly), we used the Cumby-Huizinga test to test for autocorrelation in lags. The type I error rate was 0.05 and all tests were two-sided.

Weekly models:

We chose to implement weekly models as a sensitivity check for the main daily model effect estimates. Using population denominators from the South African Community Survey 2016 (provided by IPUMS International), we calculated the average number of daily first doses administered per 10,000 individuals by week for the ≥60 years old population at national and provincial levels for weeks beginning on Monday, August 2, 2021, through November 27 2021. Based on visual inspection, we began pre-intervention data for weekly models on August 16th, 2021. (eFigure 2). Like the daily models, national weekly models were implemented using linear regression with Newey-West standard errors, and province-level models were implemented using generalized estimating equations with robust standard errors. National-level models did not include any other covariates, while province-level models included an indicator for the provinces of KwaZulu-Natal and Gauteng compared to the rest of the country, as well as an interaction term for the impact of the program in KwaZulu-Natal and Gauteng compared to the rest of the country (eTable).

**eFigure:** Average daily first dose vaccination rate per 10,000 adults ≥60 years in South Africa by week, for the weeks of August 16, 2021 through November 22, 2021

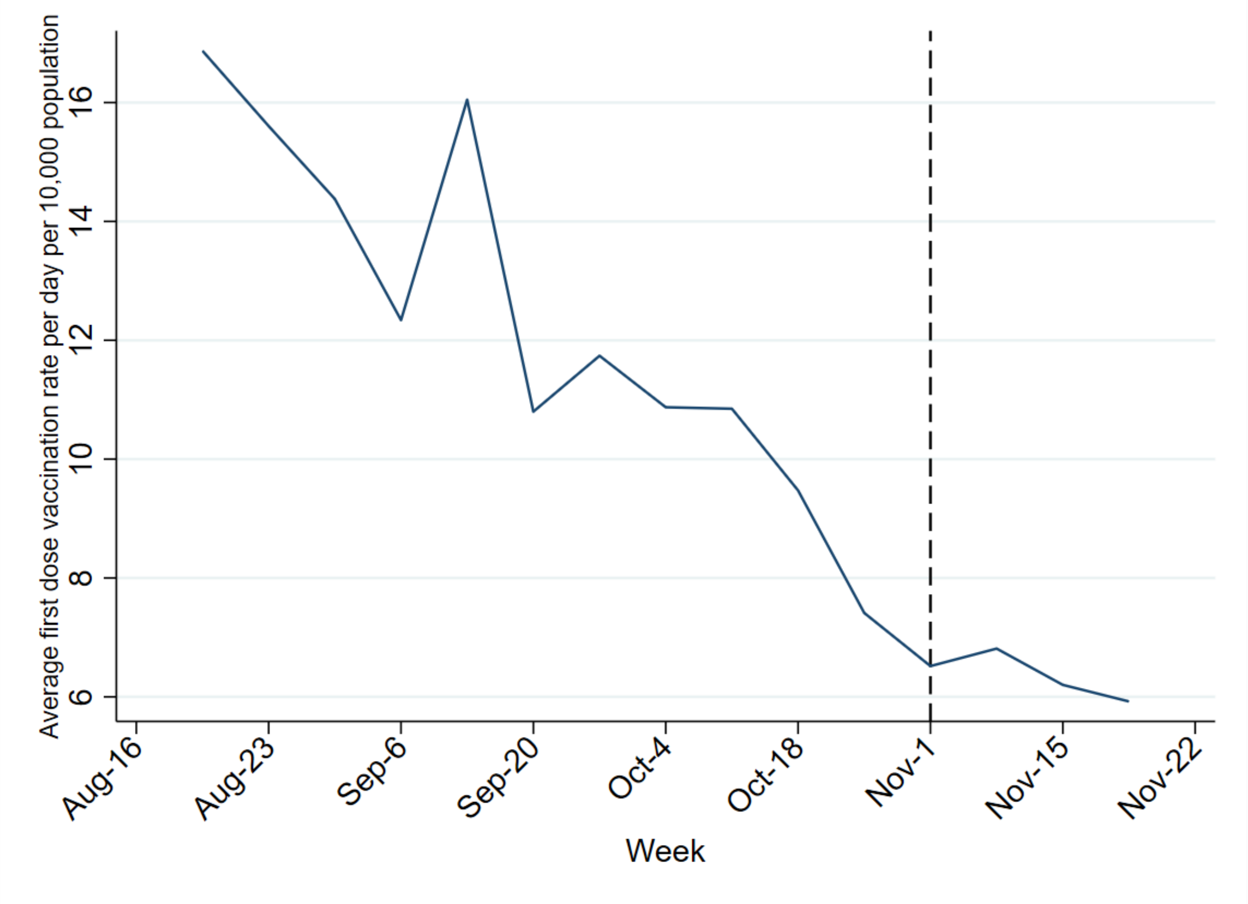

**eTable:** Results from interrupted time-series analyses estimating the association of the Vooma voucher program with changes in the average daily number of first vaccine doses delivered per week per 10,000 adults ≥60 years in South Africa

| | Unadjusted weekly models | |
|---|---|---|
| | Coeff (95% CI) | p-value |
| **National** | | |
| Vooma voucher programme | 0.58 (0.32, 0.84) | <0.001 |
| **Provincial** | | |
| Vooma voucher programme | 0.68 (0.19, 1.16) | p=0.007 |
| KZN/Gauteng | -4.27 (-6.61, -1.93) | p<0.001 |
| Voucher in KZN/Gauteng | 2.07 (1.27, 2.87) | p<0.001 |
